# Interest-holder perspectives on integrating Doing What Matters in Times of Stress with a community-based physical activity service for people experiencing mental health challenges

**DOI:** 10.64898/2026.06.24.26356509

**Authors:** Chiara Mastrogiovanni, Simon Rosenbaum, Grace McKeon, Uzma Choudhry, Sevasti Tefa, Oscar Lederman, Kemi Wright, Scott B. Teasdale, Davy Vancampfort, Gülşah Kurt

**Author notes:** Corresponding author: Professor Simon Rosenbaum. Nutrition, Exercise & Social Equity (NExuS) Research Group, Discipline of Psychiatry and Mental Health, School of Clinical Medicine, University of New South Wales Kensington, NSW 2052, Australia.

## Abstract

People experiencing mental health problems often encounter fragmented systems of care in which physical and mental health needs are addressed separately. Physical activity is an evidence-based approach for improving both physical and mental health and integrating evidence-based psychosocial support with physical activity in community settings may offer a holistic and accessible approach. This study explored interest-holder perspectives on integrating the World Health Organization’s Doing What Matters in Times of Stress intervention within a trauma-informed, community-based physical activity service. A qualitative study was conducted within a free, community-based, university-run physical activity service in Sydney, Australia. Semi-structured interviews were undertaken with people with lived expertise of mental health challenges, Clinical Exercise Professionals, and mental health service providers. Data were analysed using thematic analysis guided by the Consolidated Framework for Implementation Research. Nineteen participants (11 people with lived expertise, four Clinical Exercise Professionals, and four service providers) took part. Participants generally viewed the future delivery of Doing What Matters in Times of Stress by exercise professionals as acceptable and potentially beneficial for supporting both mental and physical health. Existing rapport with exercise professionals, the “disarming” nature of physical activity, and practical stress-management strategies were identified as strengths of the model of future delivery. Participants viewed Clinical Exercise Professionals as potentially well-placed to facilitate Doing What Matters in Times of Stress alongside supervised physical activity, as long as it was supported by appropriate training, supervision, referral pathways, and clear professional boundaries. Trauma-informed, inclusive environments, tailoring the intervention, prioritizing service user choice and organisational support were also considered important factors for successful future implementation.

**Conclusions:** Integrating Doing What Matters in Times of Stress within a trusted, community-based physical activity service was perceived as acceptable and potentially meaningful for people experiencing mental health challenges. Findings warrant further piloting and evaluation of integrated physical activity and psychosocial intervention models.

## INTRODUCTION

People living with mental health conditions are at increased risk of physical health comorbidities and premature mortality (1). An estimated 418 million disability-adjusted life years (DALYs) are attributed to poor mental health, representing 16% of global disease burden (2). Despite this significant health burden, access to health care is often limited by financial constraints, service wait times, transportation barriers. Fragmented models of care can further reduce access to appropriate support by treating mental and physical health needs separately (3, 4). There is increasing recognition that integrated approaches are needed to address mental and physical health needs concurrently to improve overall health and wellbeing of people living with mental health conditions (5, 6).

Combining evidence-based mental and physical health interventions may represent one approach to addressing the complex and interconnected health needs experienced by people facing poor mental health. One such mental health intervention is Doing What Matters in Times of Stress (DWM), developed by the World Health Organization (WHO) (7). DWM is an evidence-based, transdiagnostic stress management intervention adopting principles of acceptance and commitment therapy that provides individuals with strategies to cope with stress (7). Designed to increase accessibility, DWM is a five-week self-guided program consisting of an illustrated booklet and accompanying audio recordings that support grounding, value clarification, and self-compassion practices for people experiencing low to moderate distress. DWM can be facilitated by non-mental health specialists who are trained as DWM ‘helpers’ as the intervention can be provided with minimal guidance, facilitators mainly assisting with engagement, or with no guidance, relying on self-directed use. DWM has been translated into more than 25 languages, demonstrating effectiveness in reducing psychological distress, and improving wellbeing and functioning across a range of populations and settings (8–12).

Physical activity is recognised as an effective, acceptable, and scalable health-promotion strategy with established benefits for both physical and mental health (13, 14). Meta-analytic evidence demonstrates that physical activity can improve symptoms of depression, anxiety, and post-traumatic stress, while also supporting physical health, functioning, and quality of life across clinical and general populations (15–19). While DWM primarily aims to support psychological coping and emotional wellbeing, physical activity may provide complementary benefits through movement, behavioural activation, social connection, and improvements in physical health. However, these interventions are typically delivered separately with DWM used in mental health and psychosocial support contexts (8, 9), whereas physical activity interventions are commonly implemented through physical health, rehabilitation or community-based service pathways, often by clinical exercise professionals (CEPs) (20). Evidence suggests that integrating psychological interventions with physical activity may enhance therapeutic outcomes, although implementation considerations such as sequencing, context, and safe delivery remain important (21). Building on the strengths of both approaches, integrating physical activity programs with a brief, scalable psychological intervention may represent a feasible and accessible approach to support overall health and wellbeing among people experiencing mental health challenges.

Community-based services are locally embedded, designed and delivered in partnership with community members, to address social, mental and physical health needs in ways that are accessible, culturally responsive, and shaped by the lived realities of the people it serves (22). Community-based services may represent a trusted, accessible and less stigmatising platform for delivering integrated interventions, particularly for people with poor mental health who may already engage with these services for practical, social, or welfare support. Addi Moves, the setting of this study, is an example of a community-based physical activity service, overseen by UNSW’s Discipline of Psychiatry and Mental Health, designed to promote both physical activity and social participation among people experiencing mental health challenges (23). Located within the Addison Road Community Centre in Sydney, Australia, Addi Moves is a free, co-designed, culturally inclusive, and trauma-informed community physical activity service delivered by Accredited Exercise Physiologists (23–25). CEPs such as Accredited Exercise Physiologists are tertiary qualified allied health practitioners within the Australian healthcare system, with expertise in using physical activity in the management of chronic health conditions such as mental health challenges. This setting therefore represents a promising context to explore the implementation considerations around an intervention delivered by CEPs as non-specialist mental health workers, including an evidence based psychological intervention and physical activity sessions.

This study explored the perspectives of people with lived expertise of mental health challenges (PWLE), CEPs, and mental health service providers on the acceptability and implementation of integrating DWM with a trauma-informed, community-based physical activity service for people experiencing mental health challenges. Guided by the Consolidated Framework for Implementation Research (CFIR) (26), the study aimed to identify factors perceived to influence the acceptability, feasibility, and implementation of the integrated intervention across contextual, service, delivery, and workforce domains.

## METHODS

### Study design

This study is a qualitative, cross-sectional study. Online semi-structured interviews were conducted between September 2025 and March 2026 with some taking place physically at Addi Moves. Participants were recruited through an existing trauma-informed, community-based physical activity service (Addi Moves) and a private physical activity service, specifically, an exercise physiology clinic. Ethical approval was obtained from the University of New South Wales, Human Research Ethics Committee (iRECS5757). The reporting of this study was guided by the Consolidated criteria for Reporting Qualitative research (COREQ) (27). The full COREQ checklist can be found in the Supplementary Materials.

### Participants

19 participants were recruited from three interest-holder groups to explore perspectives on the delivery of an integrated physical activity and scalable mental health intervention delivered by non-specialist mental health practitioners. These groups included participants utilising Addi Moves physical activity service, CEPs who work with people experiencing mental health problems, and mental health support providers who could provide insights into referral pathways, client suitability, and service-level enablers and barriers.

#### 1. People with Lived Expertise of Mental Health Challenges

The Inclusion criteria for people experiencing mental health problems were being at least 18 years and having used the Addi Moves physical activity service. No exclusion was placed on number of sessions or length of engagement with Addi Moves. Addi Moves practitioners shared the study invitation with clients of Addi Moves via client email/phone details. All approached participants agreed to participate with the exception of one who did not respond.

#### 2. Clinical Exercise Professionals

CEPs self-reported as currently delivering services to clients with diagnosed mental illness or mental health challenges, were recruited through an existing trauma-informed physical activity service with exercise physiologists as facilitators (Addi Moves) and a physical activity rehabilitation service in New South Wales. No limits were placed on years of experience. Dissemination to CEPs was through broad channels, therefore the reach of the invitation was unknown and response rate could not be determined.

#### 3. Mental Health Service Providers

Mental health service providers currently delivering services to clients with diagnosed mental illness or mental health challenges and who had referred to Addi Moves, were eligible for inclusion. No limits were placed on qualification or years of experience. Addi Moves practitioners contacted service providers to inform them of the study. All approached participants agreed to participate with the exception of two who did not respond.

### Data Collection

Participants provided written informed consent before taking part in 20-30 minute semi-structured interview with the lead researchers (CM & GK). Data collection ended when data adequacy was reached, whereby interim data analysis indicated that richness and relevance of data were satisfactory based on the study aim (28). This was supported by the information power of the sample, as participants were purposively recruited for their specific experience and expertise relevant to the research question (29). The interview schedule can be found in the Supplementary Materials. Interview questions were guided by the WHO ExpandNet Framework (30) to understand interest-holder perspectives of an integrated intervention. Sessions were recorded on Microsoft Teams and auto-transcribed, then checked by the research team to check transcription was verbatim, however participant checking was not completed. Written field notes were taken throughout sessions. Eleven individual interviews, two dyadic interviews involving two participants each, and one focus group of four people were conducted with the mode based on participant preference.

### Data Analysis

Thematic analysis combining inductive and deductive approaches was used to analyse the data (31, 32). A deductive thematic analysis was conducted using the CFIR (26). Data were mapped onto relevant CFIR domains and constructs. Within each domain, inductive coding was also used to capture context-specific themes that were not fully represented within the CFIR framework. CFIR provides a comprehensive framework for examining multilevel determinants of implementation across five domains including intervention characteristics, inner setting, outer setting, characteristics of individuals, and implementation processes (26). This framework also supports the identification of practical strategies to enhance intervention adoption, feasibility, acceptability, and long-term sustainability (26).

The lead researcher (CM) first became familiar with the data by reading transcripts multiple times prior to coding. Initial, semantic codes were generated and organised according to relevant CFIR domains and constructs. Two other researchers (GK & UC) independently coded 30% of transcripts. Codes were then reviewed and refined into themes and subthemes reflecting factors influencing the acceptability and potential implementation process of the integrated intervention. Lastly, authors engaged in critical discussions to review and refine themes to ensure they accurately represented participant perspectives and the broader context of implementation. A qualitative, descriptive methodology was used (33), informed by a pragmatic research paradigm due to the practice-oriented study aims (34).

### Research Positioning

This research was conducted by a multidisciplinary team with backgrounds in exercise physiology and psychology, with experience across physical activity, mental health, and community-based service delivery. Several authors have worked directly within community organisations supporting people experiencing social disadvantage and poor mental health, which informed the practice-oriented and implementation-focused nature of this study. Additionally, authors (CM, UC, ST and OL) previously worked as Addi Moves’ practitioners and in particular, lead author (CM) who led interviewers, was known prior to some of the participants. This may have introduced social desirability bias, whereby clients felt inclined to provide more positive feedback regarding their experiences. However, the existing rapport may also have facilitated participant comfort and openness, enabling richer and more in-depth responses. Out of all the listed authors, 60% identified themselves as female and 70% held PhDs, while an additional 20% were higher degree research candidates at the time of the study.

This paper is written primarily for mental health service providers, CEPs and community organisations interested in accessible, trauma-informed, and community-based approaches to supporting mental health and wellbeing. In presenting this work, we acknowledge that the integration of physical activity and psychosocial approaches requires careful consideration of context, workforce capability, and the needs of PWLE, and should not be interpreted as suggesting that all exercise professionals are equipped to deliver such interventions without clear scope of practice, appropriate training, support, and interdisciplinary collaboration.

## RESULTS

The sample comprised 19 participants (11 PWLE, four CEPs and four mental health service providers). PWLE were aged between 30-74 years old, CEPs’ ages ranged between 24-35 years and service providers were between 40-70 years. All CEPs were had experience working with PWLE in Australia, qualified as Accredited Exercise Physiologists, while mental health service providers included two counsellors, one peer worker and one nutritionist from a mental health service that had referred PWLE to Addi Moves. A guided by the CFIR, results were summised into the ‘Integrated Intervention’, ‘Outer Setting’, ‘Inner Setting’, ‘Individuals’, and the ‘Implementation Process’.

### Characteristics of the Integrated Intervention (CFIR domain one)

All participants were supportive of an integrated physical activity and mental health intervention, alongside key implementation considerations for future delivery. Four subthemes arose in relation to CFIR domain one.

#### Multicomponent, Holistic Approach

Participants valued the two intervention components separately and welcomed an intervention that could promote both mental health and physical health in PWLE delivered by CEPs. In particular, existing rapport between CEPs and PWLE and the ability of physical activity to be ‘disarming’, were all seen as advantageous in terms of benefitting from this integrated intervention. Both service providers and CEPs also believed that their clients would mostly respond positively to the delivery of DWM alongside with physical activity.

#### Structured Manual in Various Languages

Exercise professionals expressed that they already incorporate mental health strategies such as breathing control and mindfulness into physical activity sessions however the availability of a structured DWM manual was viewed as providing clear guidance and enhancing confidence in delivering the intervention.

> *“I know a few EPs who also do box breathing, things like that. Whereas something like this, I would say is like a bit more structured. There’s something to follow and I think that’s really new. So I think that if more EPs were comfortable or sort of knew that it could be within their role or their scope to deliver things like this, I think that would really help. Alongside then being trained in how to sort of talk about it or deliver it, the confidence in the person who’s talking is always seen instantly.”* – CEP
>
> *“[The DWM manual and practice would be] similar to how a home exercise programme could work. If you give someone a booklet of exercises, they might keep it at home, but not use it. But after you’ve explained it in person, and most of the times the person gives it a go, they’re more likely to do it again. And of course, those like regular check-ins reminds them.”* – CEP

Materials translated into various languages was seen as essential to promote participation across diverse cultural and linguistic groups while visuals in materials and a booklet to take home were seen as potential facilitators to engagement.

#### Adaptability

Interest-holders emphasised the need for any integrated intervention to be flexible depending on participant needs. Delivery format was one aspect where opinions differed, supporting the need for adaptability. When discussing with PWLE the delivery format of DWM sessions, most preferred the idea of individual or small group, in person sessions with online options once rapport was built and for the purpose of maintaining momentum when an in-person session is not possible.

> *“if it’s less people, they can ask questions. But if it’s a big group, maybe it’s difficult to have your voice heard”* – PWLE

Opinions were mixed about whether the DWM content should be delivered before or after physical activity sessions, although discussions supporting that physical activity can be disarming, especially when mental health stigma is a barrier to seeking support, suggested facilitating DWM could follow a physical activity session.

#### Reminder Messages

Reminder messages were discussed as a potential facilitator for engagement. Some interest-holders suggested texts or calls to remind participants of sessions, while ideas also included using messages to promote practice of home-based physical exercise and stress-management skills.

### Healthcare Context (CFIR domain two: Outer setting)

The outer setting domain prompted discussion around one subtheme, *Partnerships and Connections*.

#### Partnerships & Connections

Interviews with mental health service providers demonstrated overarching support from referring partners for a community physical activity service to expand offerings to include stress-management strategies. Further, it was expected that physical activity services should provide physical, mental and social benefits, and this potential intervention could further the benefits that physical activity already provides. Partners shared an understanding that this intervention would remain aligned with collaborative client care.

> *“I work with survivors of child sexual abuse, I expect that there would be some kind of activity [at Addi Moves] that would help them with empowerment through embodiment. So they often have chronic pain as well. So I expect that there would be, that the exercise physiologist has that expertise to work with chronic pain and for therefore to be outcomes.”* – Service Provider

Aligned with the importance of partnership, PWLE expressed the benefit of when services are co-located such as Addi Moves’ location within a community centre providing other psychosocial supports.

> *“What makes [Addi Moves] successful is actually a constellation of things. It’s a weekly regular program, you know on a particular time, it’s a group program, there’s a positive and upbeat facilitator. We can also go to the food pantry. So, we like coming to this service. You know, we know people who have studios and stuff here. So the location is a big factor for us. If the location changed that would be one less factor. So there’s a whole range of factors that go to making it work.”* – PWLE

### The Addi Moves Physical Activity Service (CFIR domain three: Inner setting)

The inner setting in this context referred to the Addi Moves physical activity service. Under this domain, three subthemes were included.

#### Culture

Workforce culture among both CEPs and service providers was positive about the role of CEPs in mental healthcare but also to deliver holistic health interventions that includes stress management strategies other than physical activity. Participants shared an openness to the integrated approach if there is an opportunity to improve services and outcomes for PWLE.

#### Structural Characteristics

PWLE shared that previous past experiences with traditional exercise services have often made them feel uncomfortable and unsupported, while a co-designed, inclusive and trauma-informed space offers a non-judgemental space without pressure to perform or comparison. Service providers echoed the importance and expectation of a safe space for their referred PWLE to feel welcomed and empowered.

> *“I feel like at a normal gym like I I just won’t go to them because I don’t feel safe or comfortable. And I knew that I could go at my own at my own pace [at Addi Moves] and that would be supported here because we are coming in here with mental health challenges.”* – PWLE
>
> *“When I send a client, my expectation will always be that the client will be provided a safe place that they can learn and enjoy physical activity and plan short-term goals so that they can focus on the intervention, which means it will bring very good outcomes.”* – Service Provider

#### Organisational Support

Participants were generally supportive of exercise physiologists facilitating DWM as a structured, evidence-based intervention, provided delivery was supported by appropriate training, supervision, referral pathways and clear professional boundaries. However, it was expected that CEPs would have the skills to practice with a trauma-informed approach. Participants also expected that organisations would offer training to CEPs in DWM facilitation. Participants who were CEPs reinforced this by sharing that training offered would allow them to feel competent to deliver DWM and professional supervision could be an opportunity for ongoing assistance.

> *“Training and resources, could be good to run through a few practise sessions with other people. And then I think further resourcing in terms of the evidence for it to sell it further would be helpful.”* – CEP

### Individuals

Participants discussed considerations around two stakeholder groups that would be involved in future delivery of this intervention. These included CEPs and service users.

#### CEPs as Facilitators

The integrated intervention introduced a novel role for CEPs as DWM ‘helpers’ or facilitators alongside their usual delivery of physical activity services. PWLE prioritised the facilitator being their trusted exercise professional and saw this opportunity as a chance to build on their existing rapport with exercise practitioners. PWLE and service providers all saw delivery of a non-specialist mental health program to be within the scope of tertiary qualified CEPs.

PWLE and service providers supported the CEP skillset as being appropriate to deliver the integrated intervention due to knowledge and experience with holistic health beyond exercise service delivery.

> *“Do you feel like there’s any concerns with exercise physiologists delivering sort of education about grounding, breathing, mindfulness?” – Interviewer*
>
> *“I don’t think so. I’m sure they should be specialised in holistic health care.”* – Service Provider

Further, participants expressed that it was crucial exercise professionals to have capacity to create a safe environment that empowered participants’ preferences on whether or not to participate and which components to engage.

> *“I think the connection is good. Even getting familiar with you guys and forge a connection, a safe place. You know, someone familiar. Sometimes there’s stuff going on, sometimes we chat about that. It’s been something to structure my day as well.”* – PWLE

Exercise professionals contributed that individual competence to deliver the intervention is required, linking to organisational support and training.

> *“I think that if more EPs were comfortable or sort of knew that it could be within their role or their scope to deliver things like [DWM], I think that would really help*.
>
> *Alongside then being trained in how to sort of talk about it or deliver it, the confidence in the person who’s talking is always seen instantly.”* – CEP

#### Defining Target Service Users

Strong support for the integrated intervention came with the caveat that the target population would need to be clearly defined as suitability may not apply universally. One example that frequently arose was that the intervention may not be suitable for people already receiving mental health support or with an extensive background of engaging with mental health treatment, aligning with DWM’s target of being for people experiencing low to moderate levels of psychological distress rather than severe mental health challenges.

> *“I’ve been doing therapy and psychoeducation for decades and something like [DWM]. It’s just a little basic.”* – PWLE

### Implementation Process

In regards to the implementation of the integrated physical activity and mental health intervention, three subthemes were considered.

#### Trusted Relationship between Participants and Practitioners

Regarding choice of facilitator, it was clear that the trusted relationship between PWLE and CEPs was valued and seen as the foundation of successful program implementation. PWLE in this study, all given the choice expressed that they would choose their exercise professional to deliver both components of the integrated intervention rather than having an additional practitioner. PWLE having existing rapport with the CEPs could support their openness to engage initially and continually.

#### Assessing Needs

Community needs and global events were discussed as influential to update and where the most benefit may be experienced. One participant suggested that the COVID-19 pandemic highlighted the value of scalable interventions that integrate self-management strategies with accessible, community-based support during periods of heightened stress and service disruption.

#### Tailoring

Participants suggested that the intervention could be tailored depending on PWLE needs, preferences and past experiences. Discussions also surrounded tailoring engagement strategies for different age groups, disability considerations such as carer availability for in-person sessions and various cultures. Using the example of DWM, participants identified tailoring was possible due to various languages of materials, the option of a booklet and/or recordings and when they chose to practice stress-management strategies. Participants, particularly PWLE emphasised that the facilitation of DWM should not interfere with or take away from the time allocated for physical activity.

> *“What are your thoughts about the potential of this integrated program?”* - Interviewer *“My initial thoughts are yes and no, which I will explain. I think it’s if someone’s coming in and they’re sort of new to physical movement, but they’re also maybe a bit newer to mental health diagnosis or the mental health world, I think it’s a great combination. I think it would work really well. But we do tend to have a lot of clients who have potentially been living with mental health problems for a very long time and sort of been in and out of psychoeducation. So I guess it’s just about tailoring it to the right people. If I look at myself, for example, I have really high levels of anxiety. And I’ve lived with anxiety for, you know, 40 years. And I know all anxiety inside and out. And if someone says to me that I just need to breathe when I’m feeling anxious, I want to slug them because I don’t find that helpful. So I say yes for the right population.”* – Service Provider
>
> *“I’m not sure if it comes in different languages, but I do have some clients that don’t speak or maybe don’t read English. So if the book is also in English, then I think that might not be very accessible for them. The other thing is probably cost. Is there any cost attached to like this sort of thing?”* – CEP
>
> *“If he goes to group sessions, it’s a little hard, because we have to organize with the support worker as well”* – PWLE

Figure 1 provides an overview of the CFIR in relation to this study.

**Figure 1.**
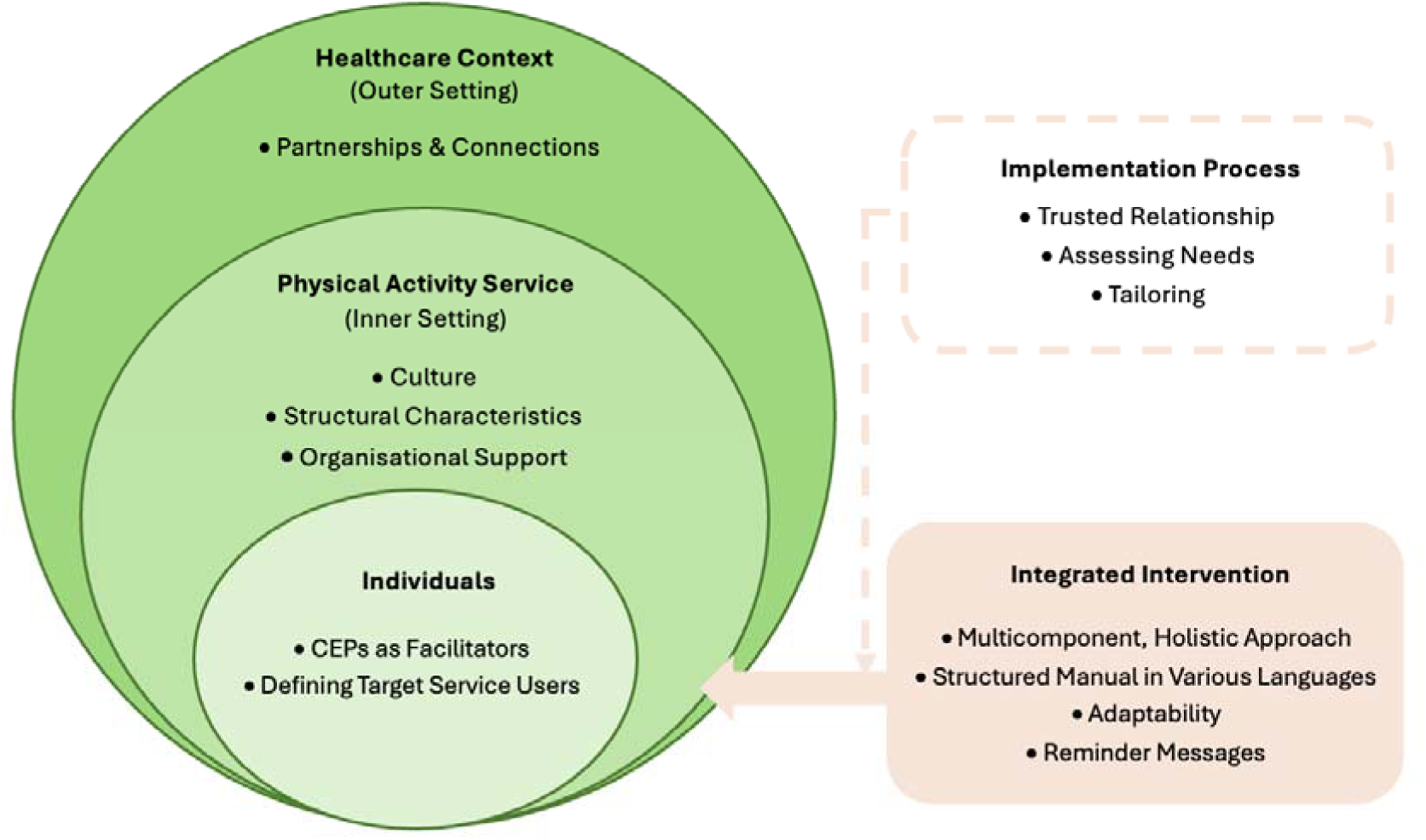
Mapping implementation factors of an integrated physical activity and mental health intervention, to the Consolidated Framework for Implementation Research.

## DISCUSSION

This study explored interest-holder perspectives on integrating Doing What Matters in Times of Stress with a trauma-informed, community-based physical activity program, delivered by Clinical Exercise Professionals to people with lived expertise of mental health challenges. Participants (PWLE, CEPs and service providers) described the integrated intervention as an acceptable, accessible, and potentially beneficial way to target both mental and physical health, particularly when delivered by a trusted practitioner. Guided by the CFIR, we identified features related to outer setting (e.g., partnerships and connections), inner settings (e.g., workforce culture and organisational support), and intervention characteristics that could support future delivery of this integrated intervention and key considerations for the characteristics of the facilitators and service users. Several important implementation processes were reported to ensure its successful delivery. Findings demonstrated the value of integrating psychosocial and movement approaches while also highlighting implementation considerations relating to exercise workforce capability, partnerships and environment. These findings suggest that CEPs may be well placed to support the use of structured, evidence-basedy psychosocial strategies such as DWM alongside physical activity, provided delivery is supported by appropriate training, supervision and interdisciplinary partnerships.

Participants were supportive of the integration of DWM and physical activity, particularly within an exercise setting perceived as safe and accessible. People with social disadvantage often experience fragmented health systems in which mental and physical health are addressed separately, despite being interrelated (3, 22). Accessible services and interventions, such as those that are community-based, offer an important opportunity to support more integrated and equitable models of care, particularly for individuals who may already engage with these settings for practical, social, or welfare support (35, 36). Participants also emphasised components shown successful in community-based physical activity services, including the importance of inclusivity, and non-judgemental environments (23, 24, 37, 38).

Participants described the integrated intervention as offering distinct but complementary benefits. DWM was perceived as providing practical strategies for emotional regulation, grounding, and coping with stress, while physical activity was associated with improvements in mood, stress management, physical health, behavioural activation, and social engagement. Importantly, participants perceived that these approaches may complement one another by simultaneously addressing various aspects of health. While psychosocial and physical activity interventions are typically delivered separately across mental health and exercise or health service pathways, findings from this study suggest that integrating these approaches within community settings may be both meaningful and acceptable for populations experiencing social disadvantage.

A key finding of this study was the perceived importance of appropriately supported delivery. While CEPs were viewed as potentially well placed to facilitate an integrated intervention due to their expertise in physical activity and chronic health conditions, interest-holders also highlighted the importance of additional training, supervision, and organisational support. Participants emphasised that workforce confidence and competence were essential to ensuring safe and appropriate delivery. This aligns with growing recognition that exercise professionals require appropriate preparation to work safely within mental health and trauma-informed contexts (25, 39–41). Importantly, these findings should not be interpreted as suggesting that all exercise professionals are equipped to deliver psychosocial interventions without additional training, support, and interdisciplinary collaboration. Rather, participants highlighted the importance of clearly defined professional roles, trauma-informed practice principles, appropriate competency development, and access to ongoing supervision and support. Discussions consistently highlighted the importance of role clarity, recognising that exercise professionals may contribute to psychosocial care through movement-based and relational approaches while working within their scope of practice and alongside mental health and other health professionals. Participants also identified structured referral pathways and interdisciplinary models of care as important mechanisms to ensure that support remains safe, ethical, and responsive to participant needs.

Findings also identified several practical and organisational considerations relevant to future implementation. Participants highlighted factors including referral pathways, accessibility, language and cultural considerations, scheduling, and organisational partnerships as important determinants of engagement and feasibility. These findings align with literature demonstrating that intervention acceptability alone is insufficient for successful implementation without consideration of contextual and organisational factors (26). The use of the CFIR helped identify factors related to intervention characteristics, workforce capability, service context, and implementation readiness that may influence future delivery. Importantly, many of these considerations reflect broader structural inequities that shape access to health-promoting opportunities among people experiencing social disadvantage (42, 43). Community-based integrated interventions may therefore require not only evidence-based intervention components but also sustained organisational support and equity-oriented implementation strategies.

### Limitations

The real-world setting increased the practical nature of findings. However, several limitations should be acknowledged. First, this study explored perspectives on a proposed delivery model rather than evaluating implementation or outcomes, therefore, findings reflect perceived rather than experienced acceptability and feasibility. Second, participants were recruited through an existing community physical activity service, and many were already engaged with Addi Moves. Their views may therefore differ from people who are not engaged in physical activity programs, have had negative experiences of exercise services, or face greater barriers to participation. Third, Addi Moves is a free, community-based and relationally oriented service, which may limit transferability to more conventional, clinical or commercial exercise settings. Fourth, participants responded to a brief description of DWM and the integrated model, rather than experiencing a fully developed intervention, training package or implementation process. As such, the study cannot determine whether exercise professionals can deliver DWM with fidelity, competence or safety, nor whether the model improves mental or physical health outcomes. Finally, the relatively small number of exercise professionals and service providers, and the likely underrepresentation of people with limited English or lower literacy, may have constrained the range of implementation perspectives captured.

## Conclusions

Overall, service users, exercise professionals and service providers viewed the integration of DWM into a trusted, community-based physical activity service as acceptable and potentially meaningful for people experiencing social disadvantage. However, acceptability was conditional on allowing PWLE’s choice, individual tailoring, trauma-informed delivery, workforce capability, referral pathways and organisational support. In order to determine feasibility, effectiveness or scalability, these findings warrant piloting the model in practice.

## AUTHORS’ CONTRIBUTIONS

The authors confirm contribution to the paper as follows: study conception and design: CM, SR and GK; data collection: CM and GK; analysis and interpretation of results: CM, UC and GK, draft manuscript preparation: CM, SR, and GK. All authors reviewed the results and approved the final version of the manuscript.

## Data Availability

Due to the qualitative nature of this study and the potential for participant identification, the full interview transcripts cannot be made publicly available. Participants were recruited from a small, specific service context (Addi Moves) and the sample included small, potentially identifiable subgroups (Clinical Exercise Professionals, n=4; mental health service providers, n=4); given these group sizes and the rich, contextual nature of qualitative data, full transcripts could risk participant re-identification even if de-identified. Participant consent and ethical approval (UNSW Human Research Ethics Committee, iREC S5757) did not extend to public deposition of interview data. Illustrative de-identified quotations supporting the study's findings are included within the manuscript. Further information about the data may be requested from the corresponding author, subject to ethics committee approval and execution of a data sharing agreement.

## ACKNOWLEDGEMENTS

The authors would like to acknowledge the support of Addison Road Community Organisation, Mindgardens Neuroscience Network and Guardian Exercise Rehabilitation.

## SUPPLEMENTARY MATERIALS

**S1.** The Consolidated criteria for Reporting Qualitative research (COREQ) checklist.

| Item No. | Topic | Description |
| --- | --- | --- |
| Domain 1: Research team and reflexivity |  |  |
| Personal characteristics |  |  |
| 1 | Interviewer/facilitator | This research was conducted by a multidisciplinary team with backgrounds in exercise physiology and psychology, with experience across physical activity, mental health, and community-based service delivery. Several authors have worked directly within community organisations supporting people experiencing social disadvantage and poor mental health, which informed the practice-oriented and implementation-focused nature of this study. Additionally, authors (CM, UC, ST and OL) previously worked as Addi Moves’ practitioners and in particular, lead author (CM) who led interviewers, was known prior to some of the participants. This may have introduced social desirability bias, whereby clients felt inclined to provide more positive feedback regarding their experiences. However, the existing rapport may also have facilitated participant comfort and openness, enabling richer and more in-depth responses. Out of all the listed authors, 60% were female and 70% held PhDs, while an additional 20% were higher degree research candidates at the time of the study. |
| 2 | Credentials |  |
| 3 | Occupation |  |
| 4 | Gender |  |
| 5 | Experience and training |  |
| Relationship with participants |  |  |
| 6 | Relationship established | Additionally, authors (CM, UC, ST and OL) previously worked as Addi Moves’ practitioners and in particular, lead author (CM) who led interviewers, was known prior to some of the participants. This may have introduced social desirability bias, whereby clients felt inclined to provide more positive feedback regarding their experiences. However, the existing rapport may also have facilitated participant comfort and openness, enabling richer and more in-depth responses. |
| 7 | Participant knowledge of the interviewer |  |
| 8 | Interviewer characteristics |  |
| Domain 2: Study design |  |  |
| Theoretical framework |  |  |
| 9 | Methodological orientation and theory | *Data Analysis*<br>Thematic analysis combining inductive and deductive approaches was used to analyse the data (31, 32). A deductive thematic analysis was conducted using the CFIR (26). Data were mapped onto relevant CFIR domains and constructs. Within each domain, inductive coding was also used to capture context-specific themes that were not fully represented within the CFIR framework. CFIR provides a comprehensive framework for examining multilevel determinants of implementation across five domains including intervention characteristics, inner setting, outer setting, characteristics of individuals, and implementation processes (26). This framework also supports the identification of practical strategies to enhance intervention adoption, feasibility, acceptability, and long-term sustainability (26).<br>The lead researcher (CM) first became familiar with the data by reading transcripts multiple times prior to coding. Initial, semantic codes were generated and organised according to relevant CFIR domains and constructs. Two other researchers (GK & UC) independently coded 30% of transcripts. Codes were then reviewed and refined into themes and subthemes reflecting factors influencing the acceptability and potential implementation |
|  |  | process of the integrated intervention. Lastly, authors engaged in critical discussions to review and refine themes to ensure they accurately represented participant perspectives and the broader context of implementation. A qualitative, descriptive methodology was used (33), informed by a pragmatic research paradigm due to the practice-oriented study aims (34). |
| **Participant selection** |  |  |
| 10 | Sampling | **Participants**<br>Nineteen participants were recruited from three interest-holder groups to explore perspectives on the delivery of an integrated physical activity and scalable mental health intervention delivered by non-specialist mental health practitioners. These groups included participants utilising Addi Moves physical activity service, CEPs who work with people experiencing mental health problems, and mental health support providers who could provide insights into referral pathways, client suitability, and service-level enablers and barriers.<br>1. People with Lived Expertise of Mental Health Challenges<br>The Inclusion criteria for people experiencing mental health problems were being at least 18 years and having used the Addi Moves physical activity service. No exclusion was placed on number of sessions or length of engagement with Addi Moves. Addi Moves practitioners shared the study invitation with clients of Addi Moves via client email/phone details. All approached participants agreed to participate with the exception of one who did not respond. |
| 11 | Method of approach |  |
| 12 | Non-participation |  |
|  |  | 2. CEPs<br>CEPs self-reported as currently delivering services to clients with diagnosed mental illness or mental health challenges, were recruited through an existing trauma-informed physical activity service with exercise physiologists as facilitators (Addi Moves) and a physical activity rehabilitation service in New South Wales. No limits were placed on years of experience. Dissemination to CEPs was through broad channels, therefore the reach of the invitation was unknown and response rate could not be determined.<br>3. Mental Health Service Providers<br>Mental health service providers currently delivering services to clients with diagnosed mental illness or mental health challenges and who had referred to Addi Moves, were eligible for inclusion. No limits were placed on qualification or years of experience. Addi Moves practitioners contacted service providers to inform them of the study. All approached participants agreed to participate with the exception of two who did not respond. |
| 13 | Sample size | The sample comprised 19 participants (11 PWLE, four CEPs and four mental health service providers). PWLE were aged between 30-74 years old, CEPs' ages ranged between 24-35 years and service providers were between 40-70 years. All CEPs were had experience working with PWLE in Australia, qualified as Accredited Exercise Physiologists, while mental health service providers included two counsellors, one peer worker and one nutritionist from a mental health service that had referred PWLE to Addi Moves. |
| **Setting** |  |  |
| 14 | Setting of data collection | Online semi-structured interviews were conducted between September 2025 and March 2026 with some sessions taking place physically at Addi Moves. |
| 15 | Presence of non-participants | N/A |
| 16 | Description of sample | The sample comprised 19 participants (11 PWLE, four CEPs and four mental health service providers). PWLE were aged between 30-74 years old, CEPs' ages ranged between 24-35 years and service providers were between 40-70 years. All CEPs were had experience working with PWLE in Australia, qualified as Accredited Exercise Physiologists, while mental health service providers included two counsellors, one peer worker and one nutritionist from a mental health service that had referred PWLE to Addi Moves. |
| **Data collection** |  |  |
| 17 | Interview guide | The interview schedule can be found in the Supplementary Materials. Interview questions were guided by the WHO ExpandNet Framework (30) to understand interest-holder perspectives of an integrated intervention. |
| 18 | Repeat interviews | N/A |
| 19 | Audio/visual recording | Sessions were recorded on Microsoft Teams. |
| 20 | Field notes | Written field notes were taken throughout sessions. |
| 21 | Duration | Participants provided written informed consent before taking part in 20-30 minute semi-structured interview with the lead researcher (CM). |
| 22 | Data saturation | Data collection ended when data adequacy was reached, whereby interim data analysis indicated that richness and relevance of data were satisfactory based on the study aim (28). This was supported by the information power of the sample, as participants were purposively recruited for their specific experience and expertise relevant to the research question (29). |
| 23 | Transcripts returned | Sessions were recorded on Microsoft Teams and auto-transcribed, then checked by the research team to check transcription was verbatim, however participant checking was not completed. |
| **Domain 3: Analysis and findings** |  |  |
| **Data analysis** |  |  |
| 24 | Number of data coders | *Data Collection* |
| 25 | Description of the coding tree | Participants provided written informed consent before taking part in 20-30 minute semi-structured interview with the lead researcher (CM). Data collection ended when data adequacy was reached, whereby interim data analysis indicated that richness and relevance of data were satisfactory based on the study aim (28). This was supported by the information power of the sample, as participants were purposively recruited for their specific experience and expertise relevant to the research question (29). The interview schedule can be found in the Supplementary Materials. Interview questions were guided by the WHO ExpandNet Framework (30) to understand interest-holder perspectives of an integrated intervention. Sessions were recorded on Microsoft Teams and auto-transcribed, then checked by the research team to check transcription was verbatim, however participant checking was not completed. Written field notes were taken throughout sessions. Eleven individual interviews, two dyadic interviews involving two participants each, and one focus group of four people were conducted with the mode based on participant preference.<br>*Data Analysis*<br>Thematic analysis combining inductive and deductive approaches was used to analyse the data (31, 32). A deductive thematic analysis was conducted using the CFIR (26). Data were mapped onto relevant CFIR domains and constructs. Within each domain, inductive coding was also used to capture context-specific themes that were not fully represented within the CFIR framework. CFIR provides a comprehensive framework for examining multilevel determinants of implementation across five domains including intervention characteristics, inner setting, outer setting, characteristics of individuals, and implementation processes (26). This framework also supports the identification of practical strategies to enhance intervention adoption, feasibility, acceptability, and long-term sustainability (26).<br>The lead researcher (CM) first became familiar with the data by reading transcripts multiple times prior to coding. Initial, semantic codes were generated and organised according to relevant CFIR domains and constructs. Two other researchers (GK & UC) independently coded 30% of transcripts. Codes were then reviewed and refined into themes and subthemes reflecting factors influencing the acceptability and potential implementation process of the integrated intervention. Lastly, authors engaged in critical discussions to review and refine themes to ensure they accurately represented participant perspectives and the broader context of implementation. A qualitative, descriptive methodology was used |
| 26 | Derivation of themes |  |
| 27 | Software |  |
| 28 | Participant checking |  |
|  |  | (33), informed by a pragmatic research paradigm due to the practice-oriented study aims (34). |
| **Reporting** |  |  |
| 29 | Quotations presented | Please see results, Figure 1 and further example quotations in Supplementary Materials. |
| 30 | Data and findings consistent |  |
| 31 | Clarity of major themes |  |
| 32 | Clarity of minor themes |  |

*Developed from: Tong A, Sainsbury P, Craig J. Consolidated criteria for reporting qualitative research (COREQ): a 32-item checklist for interviews and focus groups. International Journal for Quality in Health Care. 2007;19(6):349–357.*

## S2. PWLE Interview Schedule

1. When joining physical activity sessions with Addi Moves, what were your expectations of the service?
2. Has our service met your expectations?

a. If Yes, could you tell me how?
b. If No, could you tell me why?
3. If we were to improve our Addi Moves service, what are the things that we can do to:

a. Further meet your physical and mental health needs
b. Better use our service to help more of the community
c. Increase ease of access for yourself and others
4. At Addi Moves, we aim to improve mental health through physical activity. To improve our services, we are currently in the process of developing a 5-week program including physical activity classes and self-help stress management strategies that individuals in the program can learn to manage stress better. This program is planned to be provided by exercise physiologists who will provide weekly exercise sessions and also facilitate the use of a self-help stress management program. I will now play the brief introduction video of this self-help program. *Showing participants the Doing What Matters in Times of Stress summary video - https://www.youtube.com/watch?v=E3Cts45FNrk*
5. Would you be interested in learning about stress management (such as using this program) alongside physical activity classes?
6. If we did include stress management activities alongside our physical activity services, how would you like this delivered to you?

a. Prompt: Before, during or after physical activity sessions
b. Prompt: Group or individual sessions
c. Prompt: Online or delivered by an exercise physiologist
7. What are the things that potentially help us to deliver this combined program?
8. What might be potential barriers to service users participating in this combined program?

## S3. Exercise Physiologist Interview Schedule

1. From your perspective, what do you see as the main role of exercise physiology in supporting people living with mental health challenges?
2. What challenges do you see when delivering physical activity programs for people experiencing mental health and social disadvantage?
3. Our team are currently in the process of developing a 5-week program including physical activity classes and self-help stress management strategies that individuals in the program can learn to manage stress better. This program is planned to be provided by exercise physiologists who will provide weekly exercise sessions and also facilitate the use of a self-help stress management program. I will now play the brief introduction video of this self-help program. *Showing participants the Doing What Matters in Times of Stress summary video - https://www.youtube.com/watch?v=E3Cts45FNrk*
4. What are your initial thoughts on this idea?
5. How do you see stress management strategies fitting alongside physical activity sessions?

a. Prompt: delivered within the same session vs separate sessions
b. Prompt: group vs individual delivery
c. Prompt: online resources vs exercise physiologist-facilitated activities
6. Do you think your clients would you be interested in learning about stress management (such as using this program) alongside physical activity classes?
7. What factors would help exercise physiologists deliver this combined program successfully?
8. What barriers might exercise physiologists face in delivering this program?
9. What support, training, or resources would be most useful for you (or other EPs) if you were to deliver this type of program in the future?

## S4. Service Provider Interview Schedule

1. When you refer clients to physical activity or exercise physiology services, what are your expectations of those services?
2. Thinking about your clients, what benefits do you see in linking them with physical activity services?
3. Our team are currently in the process of developing a 5-week program including physical activity classes and self-help stress management strategies that individuals in the program can learn to manage stress better. This program is planned to be provided by exercise physiologists who will provide weekly exercise sessions and also facilitate the use of a self-help stress management program. I will now play the brief introduction video of this self-help program. *Showing participants the Doing What Matters in Times of Stress summary video - https://www.youtbe.com/watch?v=E3Cts45FNrk*
4. What are your initial thoughts on this idea?
5. Do you think your clients would you be interested in learning about stress management (such as using this program) alongside physical activity classes?
6. What would make you more likely to refer clients to this type of combined program?
7. What do you think might be barriers to your clients engaging in such a program?
8. What barriers might you or your service face in supporting referrals to this program?

---

**S5.** Full list of interview quotes organised by CFIR domains.

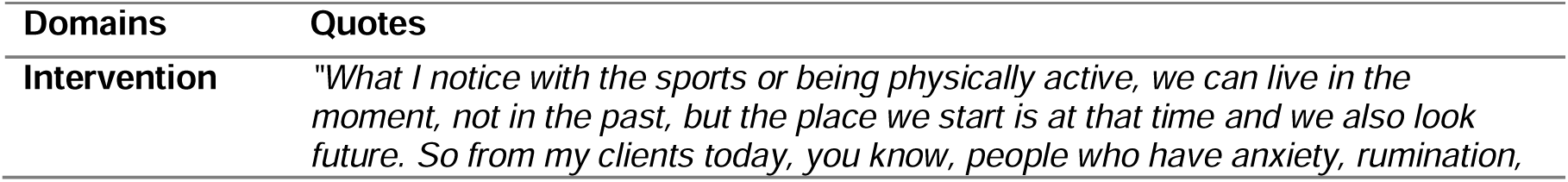
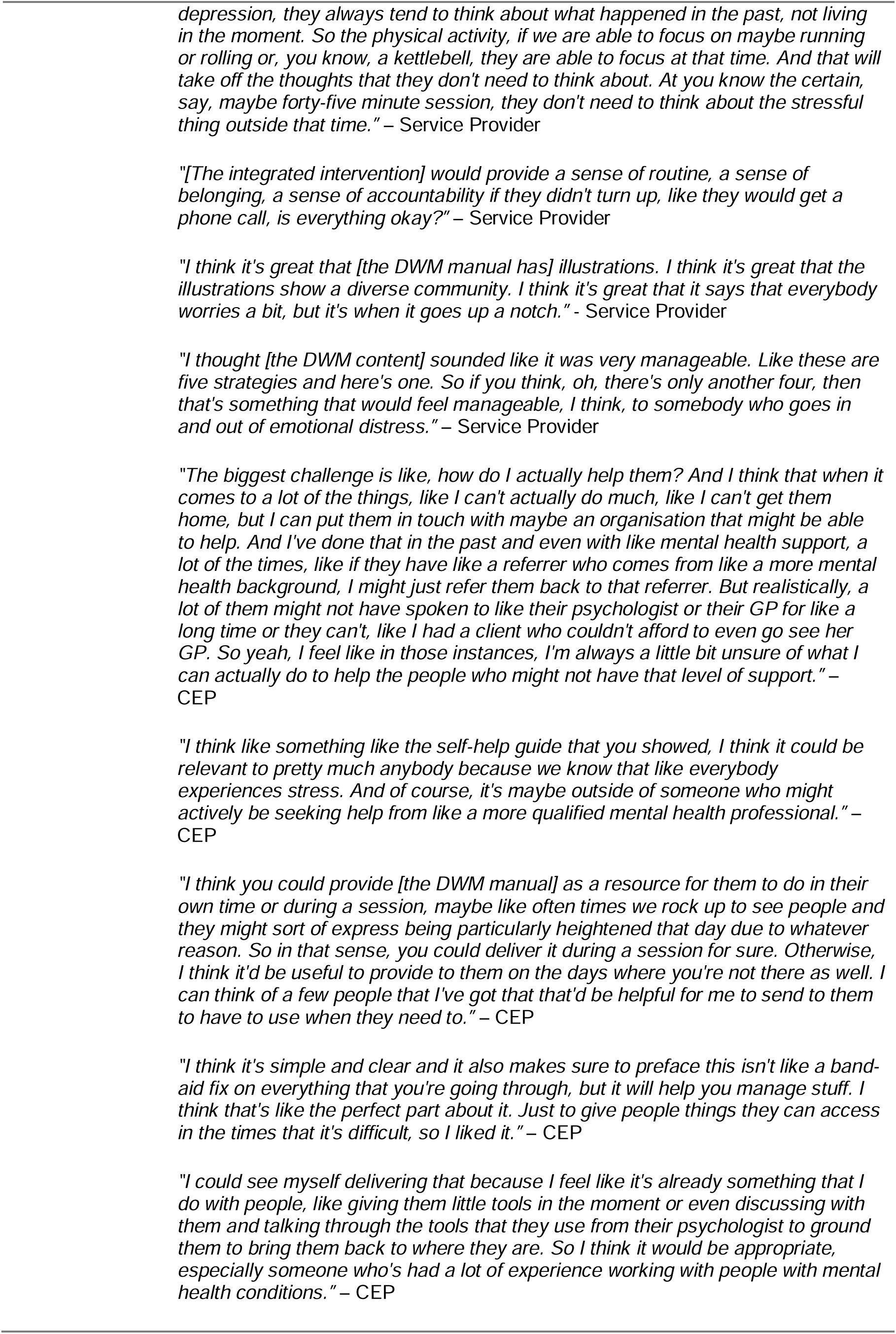
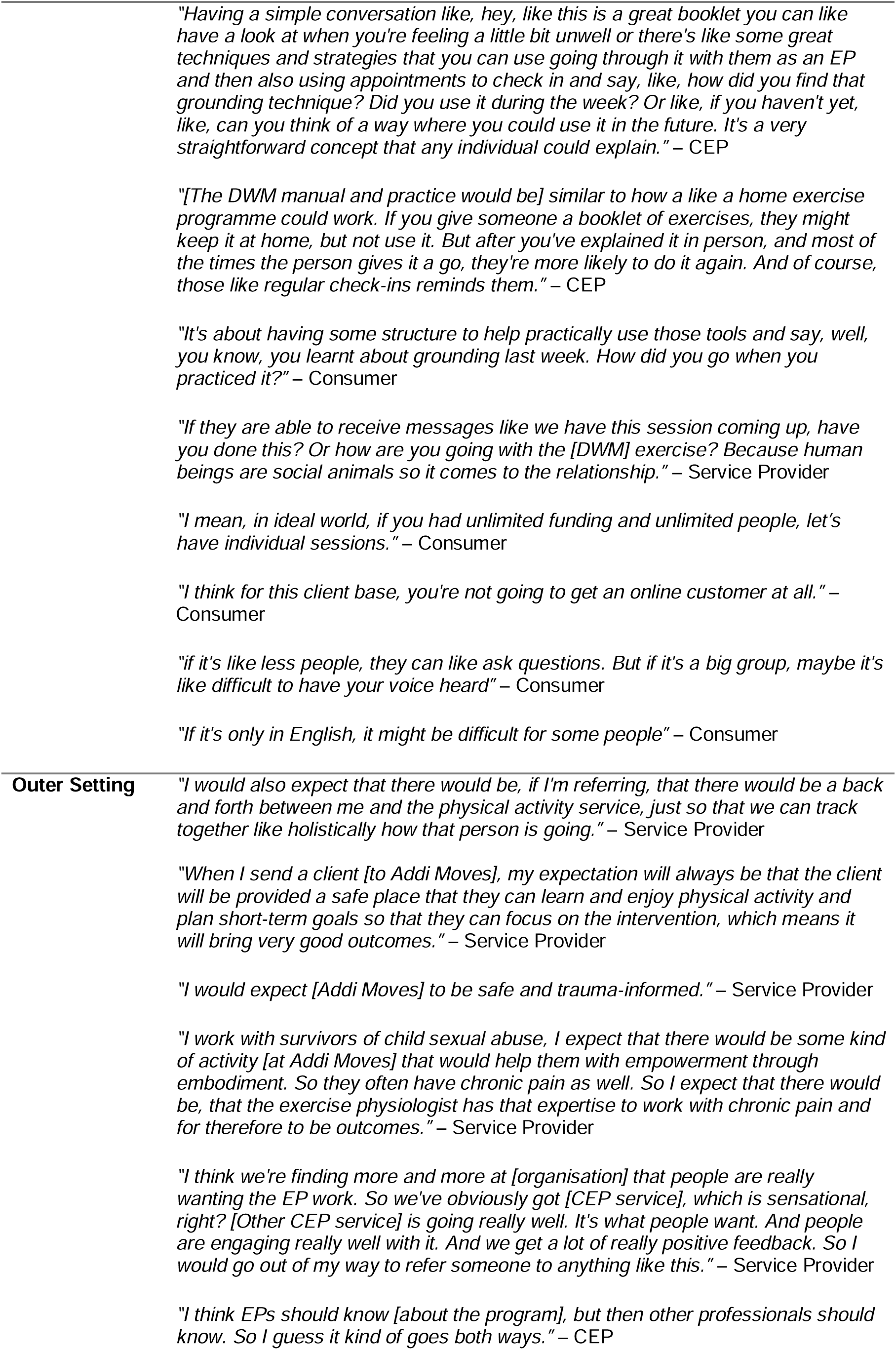
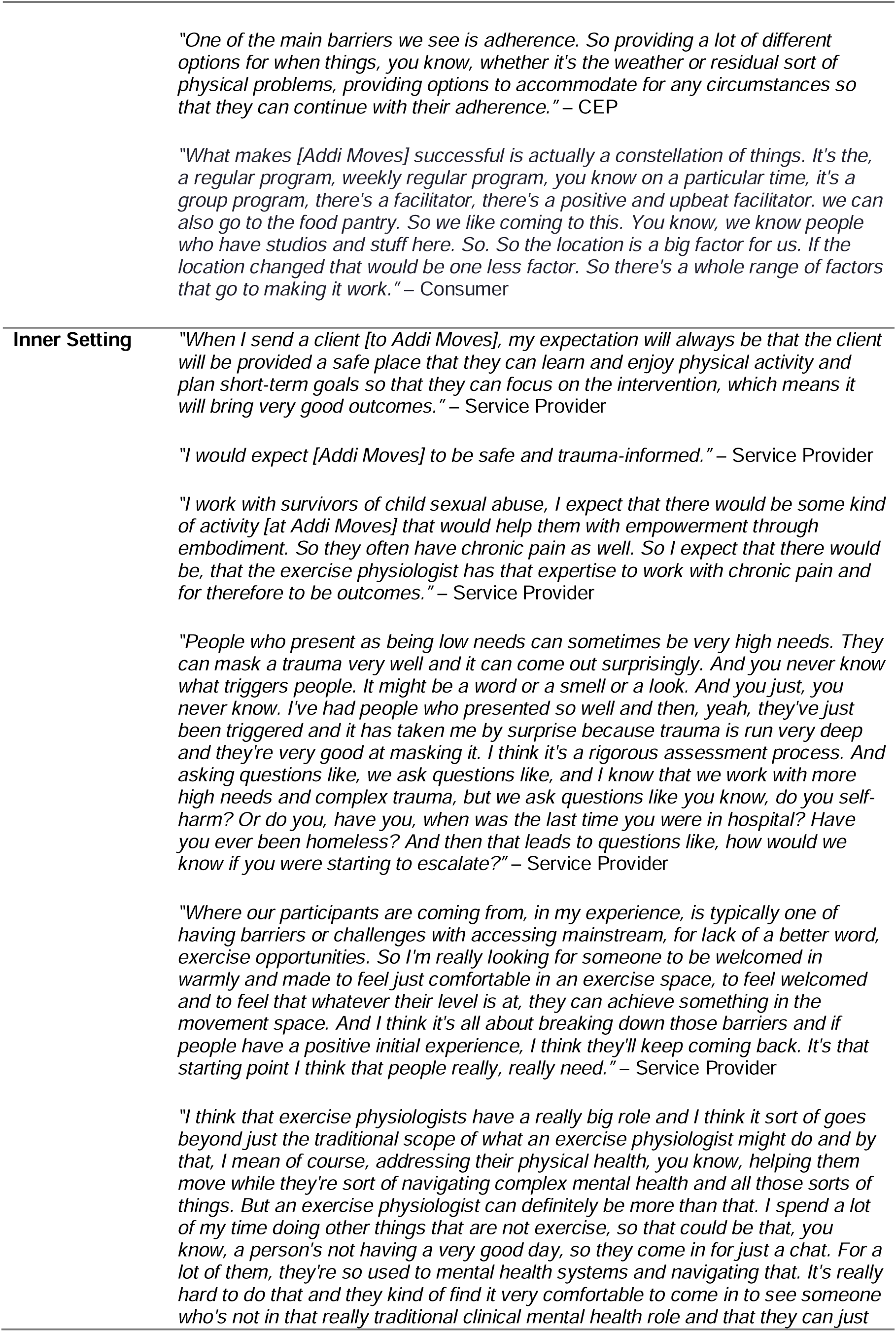
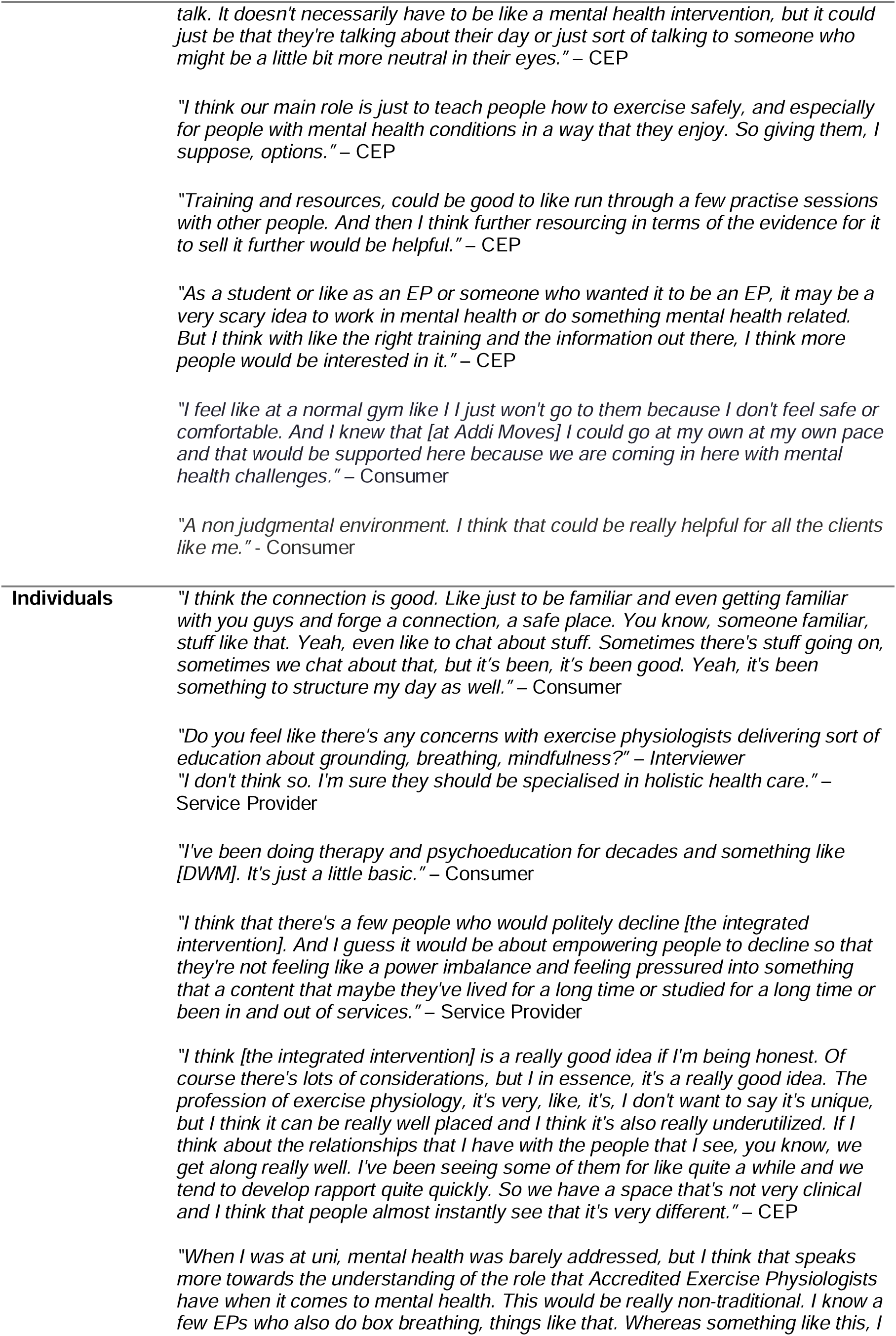
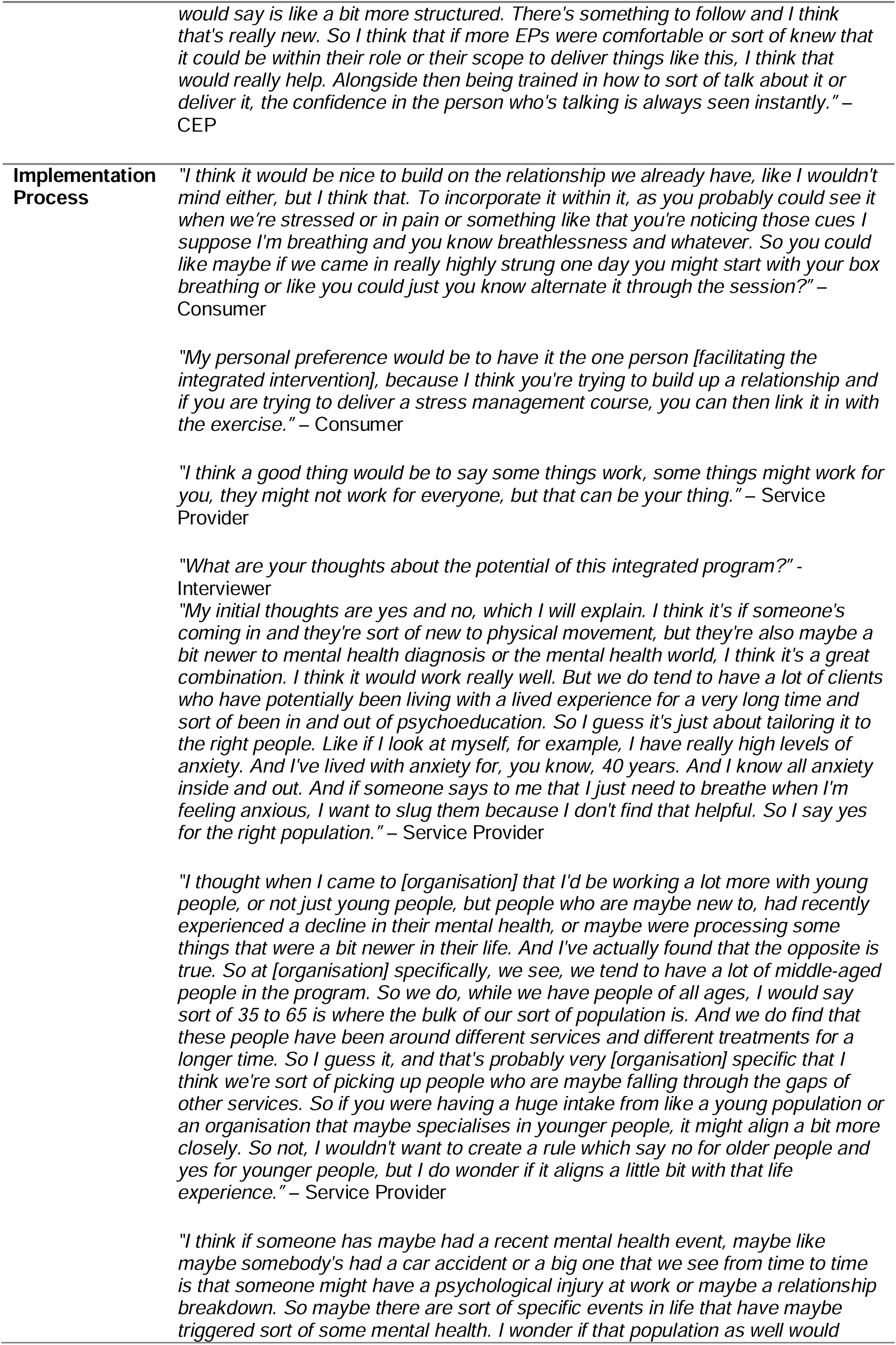
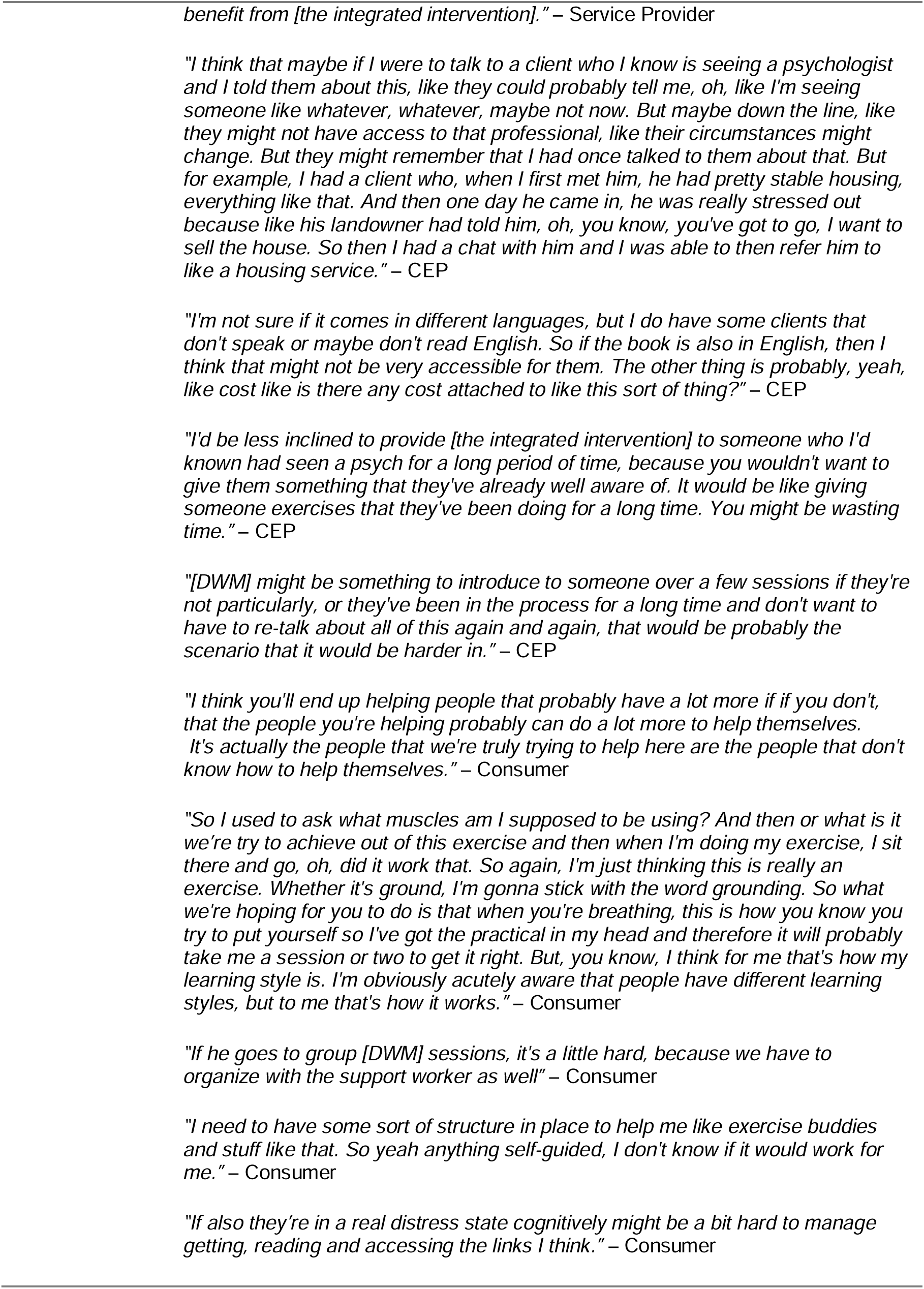

